# Circulating miRNA profiling identifies hsa-miR-23a-3p and a three-miRNA signature associated with serous ovarian cancer

**DOI:** 10.64898/2026.11.18.26368404

**Authors:** Guillem Garcia-Llorens, Ángela Martín Tierno, Anna Serra Rubert, Esther Lizano González, Antoni Llueca, Meryem El Ouardi

## Abstract

High-grade serous ovarian carcinoma (HGSOC) is the most lethal subtype of epithelial ovarian cancer, highlighting the need for minimally invasive biomarkers that support disease detection and molecular characterization. Circulating microRNAs (miRNAs) have emerged as promising candidates, although their clinical applicability remains limited by methodological variability and insufficient validation. Plasma samples from 100 individuals (50 patients with ovarian cancer and 50 cancer-free controls) were initially collected from four biobanks. Following quality control, 94 samples (47 patients and 47 controls) were included in the final analysis. Eight candidate miRNAs were quantified using RT-qPCR, and relative expression was assessed following global mean normalization. Differential expression was evaluated using the Wilcoxon rank-sum test, while discriminatory performance was assessed using receiver operating characteristic (ROC) curve analysis. Three-miRNA logistic regression models were additionally explored using repeated stratified five-fold cross-validation. Among the eight candidates, hsa-miR-23a-3p exhibited significantly lower circulating expression in patients with ovarian cancer than in controls (p = 2.14 × 10^−5^), achieving an area under the ROC curve (AUC) of 0.755 (95% CI, 0.657-0.852), with 93.6% sensitivity and 53.2% specificity at the selected threshold. Exploratory analysis across FIGO stages revealed heterogeneous expression patterns, although the limited number of early-stage cases precluded definitive conclusions regarding stage-specific performance. A three-miRNA signature comprising hsa-miR-23a-3p, hsa-miR-21-5p, and hsa-miR-20a-5p achieved an apparent AUC of 0.798. However, repeated cross-validation yielded a mean AUC of 0.716 ± 0.022, indicating reduced performance after internal validation. Variability in exogenous spike-in Ct values and the marked imbalance in biobank representation between study groups highlighted potential pre-analytical confounding. Circulating hsa-miR-23a-3p showed moderate discriminatory performance between patients with ovarian cancer and cancer-free controls, supporting its further investigation as a candidate liquid biopsy biomarker. Although combining three miRNAs improved apparent classification performance, this advantage was not established through internal validation. These findings underscore the importance of accounting for sample provenance, technical variability, and model overfitting when evaluating circulating miRNA signatures. Independent validation in appropriately balanced cohorts is required before clinical applicability can be established.

## 1. Introduction

Ovarian cancer remains one of the most lethal gynecological malignancies and represents a major challenge in women.s health worldwide. According to the latest GLOBOCAN estimates, approximately 330,731 new cases and 203,850 deaths were recorded globally in 2024 [1]. Among epithelial ovarian malignancies, high-grade serous ovarian carcinoma (HGSOC) is the most prevalent and aggressive histological subtype, accounting for approximately 70% of cases and contributing disproportionately to ovarian cancer-related mortality [2,3]. HGSOC is characterized by extensive genomic instability, frequent TP53 alterations, and defects in homologous recombination DNA repair pathways [3,4]. Moreover, accumulating histopathological and molecular evidence indicates that a substantial proportion of these tumors originate from the distal fallopian tube epithelium, supporting the contemporary classification of ovarian, fallopian tube, and primary peritoneal high-grade serous carcinomas within a common clinicopathological framework [3–5].

Despite advances in surgical management, platinum-based chemotherapy, and molecularly targeted therapies, the clinical burden of HGSOC remains substantial [6]. Most patients are diagnosed with advanced-stage disease, partly because early symptoms are nonspecific and effective population-based screening strategies remain unavailable [2,3]. Serum cancer antigen 125 (CA-125) and human epididymis protein 4 (HE4), together with transvaginal ultrasonography and clinical risk algorithms, are widely investigated or used in the evaluation of suspected ovarian malignancy. However, their diagnostic performance varies according to disease stage, histological subtype, and clinical context [7]. Importantly, long-term results from the UK Collaborative Trial of Ovarian Cancer Screening (UKCTOCS) demonstrated that screening strategies based on longitudinal CA-125 measurements or ultrasonography did not significantly reduce ovarian or fallopian tube cancer mortality [8]. These limitations underscore the need for complementary biomarkers capable of improving the molecular characterization of disease and supporting minimally invasive diagnostic approaches.

Liquid biopsy has emerged as a promising strategy for investigating tumor-associated molecular alterations through the analysis of circulating nucleic acids, extracellular vesicles, and other tumor-related components in biological fluids [9]. Among these analytes, circulating microRNAs (miRNAs) have attracted considerable attention because of their regulatory functions, detectability in blood, and potential association with tumor-related biological processes [10,11]. MiRNAs are small non-coding RNA molecules, generally approximately 18-25 nucleotides in length, that regulate gene expression predominantly through post-transcriptional mechanisms. Aberrant miRNA expression contributes to multiple hallmarks of cancer, including sustained proliferation, resistance to apoptosis, epithelial-mesenchymal transition, invasion, metastatic dissemination, and modulation of the tumor microenvironment [11]. In ovarian cancer, several miRNA families, including miR-200, miR-21, and miR-125, have been investigated in relation to tumor biology and their potential clinical applications [11].

A particularly attractive characteristic of circulating miRNAs is their relative stability in extracellular environments. Their association with extracellular vesicles, lipoprotein complexes, or RNA-binding proteins can protect them against degradation, facilitating their detection in plasma and serum [12]. Furthermore, circulating miRNA expression patterns may reflect contributions from tumor cells, stromal and immune compartments, and systemic responses to malignancy. These properties have encouraged the investigation of circulating miRNAs as candidate biomarkers for cancer detection, patient stratification, disease monitoring, and therapeutic response assessment [10–12]. Nevertheless, the biological origin of individual circulating miRNAs is not always tumor-specific, and alterations detected in peripheral blood may arise from multiple physiological or pathological processes.

Recent systematic reviews and meta-analyses have highlighted both the potential and limitations of circulating miRNAs in ovarian cancer. A comprehensive meta-analysis published in 2024 investigated their performance across different disease stages, supporting further evaluation of stage-associated circulating miRNA profiles [13]. More recently, a 2026 meta-analysis including 24 studies reported pooled sensitivity and specificity values of 0.749 and 0.748, respectively, indicating moderate diagnostic performance while identifying substantial heterogeneity related to biospecimen type and study characteristics [14]. Another systematic review published in 2026 examined circulating miRNAs and multivariable biomarker signatures across 91 studies, highlighting the frequent investigation of miR-200 family members, miR-21, miR-125b, and other candidates, as well as the potential advantages of combining molecular markers [11]. Additional evidence from exosomal miRNA studies further supports the investigation of extracellular miRNA signatures as potential diagnostic biomarkers [12]. However, these findings also emphasize that encouraging diagnostic accuracy estimates obtained in exploratory case-control studies do not necessarily translate into robust clinical performance.

Beyond distinguishing patients with ovarian cancer from cancer-free controls, an important question is whether circulating miRNA profiles are associated with clinicopathological characteristics, particularly disease stage and tumor dissemination. Such associations could provide information about the biological processes accompanying tumor progression and potentially support patient stratification. However, the interpretation of stage-associated differences requires particular caution, as early-stage HGSOC is relatively uncommon in available clinical cohorts and is frequently underrepresented in biomarker discovery studies [11,13]. Furthermore, the relationship between circulating miRNA abundance and tumor burden is not necessarily linear, given the contributions of active secretion, cellular turnover, immune responses, and tissue-specific miRNA expression. Consequently, evaluating individual miRNAs alongside multivariable signatures and available clinical information may provide a more comprehensive understanding of their potential relevance.

Despite increasing interest in circulating miRNAs, their translation into clinically applicable biomarkers remains limited by methodological heterogeneity and insufficient external validation. One of the principal challenges concerns pre-analytical variability associated with blood collection, sample processing, storage conditions, RNA extraction, and quantitative detection methods [15–17]. Importantly, plasma and serum cannot be considered interchangeable biological matrices. Blood coagulation and the associated activation or release of cellular components can alter the extracellular miRNA composition of serum, whereas plasma measurements are particularly sensitive to residual platelet contamination and centrifugation procedures [15,16]. Hemolysis represents another major source of interference because erythrocyte-derived miRNAs may substantially influence measured circulating profiles, even when sample contamination is not immediately apparent [16,17]. These factors can introduce systematic differences unrelated to tumor biology and complicate comparisons between independently collected cohorts.

Analytical normalization constitutes an additional challenge in circulating miRNA research. Unlike tissue-based gene expression studies, plasma and serum analyses lack universally accepted endogenous reference miRNAs that remain stable across diseases, sample types, and experimental conditions. Exogenous synthetic spike-in controls can help monitor technical variation during RNA extraction and downstream processing, but they do not necessarily account for biological differences in circulating RNA composition. Similarly, global mean normalization and endogenous reference-based strategies may produce different expression estimates depending on the number and stability of the miRNAs included [15]. In multicenter studies, differences in biobank procedures and sample provenance may further introduce batch-associated variation, particularly when disease status and sample origin are unevenly distributed [16,18]. Therefore, careful quality assessment, transparent normalization procedures, and appropriately designed validation strategies are essential for distinguishing potentially meaningful biological signals from technical confounding [15,19].

In this context, the present study investigated the circulating expression profiles of eight candidate miRNAs in plasma samples from patients with high-grade serous ovarian carcinoma and cancer-free controls obtained from four biobanks. Particular attention was given to hsa-miR-23a-3p, identified as a candidate associated with differences in circulating expression between study groups. We evaluated its discriminatory performance and explored its distribution according to FIGO stage. In addition, we investigated whether multivariable combinations of circulating miRNAs could improve classification performance compared with individual candidates. Given the multicenter origin of the samples, technical variability associated with exogenous spike-in measurements and the potential influence of biobank-related confounding were explicitly considered when interpreting the findings. This study therefore aims to contribute to the characterization of circulating miRNA profiles in HGSOC while highlighting the methodological considerations required for their reproducible evaluation as candidate liquid biopsy biomarkers [20].

## 2. Materials and Methods

### 2.1. Study population and plasma samples

EDTA plasma samples were collected from 50 patients diagnosed with ovarian high grade serous carcinoma (HGSOC) and 50 healthy individuals. Samples were obtained through the Biobanco del Sistema de Salud de Aragón (BSSA), Biobanco de Navarra Biomed, Biobanco Vasco and Biobanco para la Investigación Biomédica y en Salud Pública de la Comunidad Valenciana (IBSP-CV). Available clinicopathological information was retrospectively collected from biobank-associated records and pathology reports. Clinical variables included age at sample collection, histological diagnosis, primary tumor site, and FIGO stage. Additional information on serum cancer antigen 125 (CA-125) concentrations, BRCA1/2 mutational status, and homologous recombination deficiency (HRD) was recorded when available. Histopathological reports were reviewed to verify tumor classification, and molecular findings were interpreted according to the genetic testing methods and variant classifications reported by the corresponding institutions. Missing clinical or molecular information was retained as unavailable and was not imputed.

### 2.2. Plasma miRNA extraction

Plasma aliquots were thawed and centrifuged at 400 x g for 2 minutes at 4 °C to eliminate any precipitate prior to RNA extraction. RNA was then extracted using the Maxwell miRNA Plasma and Serum Isolation Kit (Promega, Cat. No. AS1680). Briefly, 400 μL plasma was lysed with 80 μL of Proteinase K and 230 μL of Lysis Buffer. Samples were purified according to the manufacturer.s protocol and eluted in 30 μL RNase-free water and stored at −80 °C until further use. To monitor extraction efficiency, 2 μL of cel-miR-39-3p (5.-phosphorylated) was added as an exogenous spike-in to each sample during the RNA isolation procedure.

### 2.3. cDNA synthesis and miR-Amp preamplification

Complementary DNA (cDNA) was synthesized using the TaqMan Advanced miRNA cDNA Synthesis Kit (Applied Biosystems, Cat. No. A25576) following the manufacturer.s protocol. For each reaction, 2 µL of RNA eluent was used as input. RNA was not denatured prior to synthesis to maintain structural integrity and prevent inhibition. The workflow consisted of four sequential enzymatic steps: (1) poly(A) tailing at 37 °C for 45 min; (2) adaptor ligation at 16 °C for 60 min to extend the 5. end; (3) reverse transcription using a universal RT primer; and (4) a miR-Amp reaction at 95 °C for 5 min followed by 14 cycles (95 °C for 3 seconds and 60 °C for 30 seconds) to uniformly amplify the cDNA pool. The resulting cDNA was diluted 1:10 in 0.1X TE buffer and stored at −20 °C until qPCR analysis.

### 2.4. Quantitative real-time PCR

Quantitative PCR (qPCR) was performed on a QuantStudio 5 Real-Time PCR System (Applied Biosystems). Reactions were carried out using TaqMan Advanced miRNA Assays and TaqMan Fast Advanced Master Mix. These assays utilize TaqMan MGB probes, which include a fluorescent reporter dye at the 5. end and a non-fluorescent quencher (NFQ) and minor groove binder (MGB) at the 3. end for enhanced specificity. Each reaction was performed in a total volume of 13.3 µL, consisting of 10 µL of PCR Reaction Mix (2X Master Mix and 20X Assay) and 3.3 µL of the 1:10 diluted cDNA template. To ensure technical consistency, all reactions were performed in triplicate, and No Template Controls (NTC) were included in every run to monitor for potential contamination or non-specific amplification. The fast-cycling mode was selected with the following conditions: enzyme activation at 95 °C for 20 sec, followed by 40 cycles of denaturation at 95 °C for 1 sec and annealing/extension at 60 °C for 20 seconds. Data were analyzed using relative quantification (2^-ΔCt^) method.

### 2.5. miRNA selection and data normalization

A panel of human miRNAs was selected for qPCR analysis based on their reported involvement in ovarian cancer and their potential utility as circulating biomarkers. Eight candidate miRNAs were evaluated: hsa-miR-23a-3p, hsa-miR-21-5p, hsa-miR-125b-5p, hsa-miR-93-5p, hsa-miR-20a-5p, hsa-miR-200c-3p, hsa-miR-200a-3p, and hsa-miR-320b. Additionally, hsa-miR-103a-3p was included as a potential endogenous reference, whereas synthetic cel-miR-39-3p (Assay ID: 478293_mir) was added as an exogenous spike-in control to monitor RNA extraction efficiency. The complete list of miRNAs and their corresponding assay identifiers is provided in Supplementary Table S1. For the exploratory expression analyses, candidate miRNA Ct values were normalized using the global mean Ct of the eight candidate miRNAs measured in each sample. Relative expression was calculated as 2^-ΔCt^, where ΔCt represents the difference between the Ct of the target miRNA and the sample-specific global mean Ct. This normalization approach was applied consistently across samples.

### 2.6. Statistical analysis

Statistical analyses were performed using R statistical software. Continuous variables were summarized as mean ± standard deviation (SD) or median and interquartile range (IQR), as appropriate, whereas categorical variables were expressed as frequencies and percentages. Relative circulating miRNA expression was calculated as 2^−ΔCt^, where ΔCt was obtained by subtracting the sample-specific global mean Ct of the eight candidate miRNAs from the Ct of each target miRNA. Differences in circulating miRNA expression between patients with high-grade serous ovarian carcinoma (HGSOC) and controls were evaluated using two-sided Wilcoxon rank-sum tests. Associations between circulating miRNA levels and FIGO stage were explored descriptively, including comparisons between early-stage (FIGO I-II) and advanced-stage (FIGO III-IV) disease. Given the limited number of early-stage cases, stage-related analyses were considered exploratory. The discriminatory performance of individual circulating miRNAs was assessed using receiver operating characteristic (ROC) curve analysis. Areas under the ROC curves (AUCs) and their 95% confidence intervals (CIs) were calculated using the pROC package, with confidence intervals estimated using DeLong.s method. Sensitivity and specificity were determined at the threshold maximizing the Youden index. To investigate the potential discriminatory value of combined circulating miRNAs, multivariable logistic regression models were constructed using normalized ΔCt values. All 56 possible three-miRNA combinations were evaluated, and the combination achieving the highest apparent AUC was identified. To assess the robustness of this model-selection strategy, repeated stratified five-fold cross-validation was performed over 20 repetitions, with miRNA combination selection repeated within each training partition. Cross-validated AUC values were summarized as mean ± SD across repetitions. Apparent and cross-validated performance estimates were distinguished to account for potential model-selection optimism. All statistical tests were two-sided, and p-values below 0.05 were considered statistically significant. Given the exploratory nature of the study and the evaluation of multiple candidate miRNAs, results were interpreted cautiously, particularly in the absence of independent validation. The marked imbalance in biobank representation between study groups was considered a potential source of confounding. The association between circulating hsa-miR-23a-3p relative expression (2^−ΔCt^) and serum CA-125 concentrations was evaluated using Spearman.s rank correlation coefficient (ρ) in the subset of 17 patients with available CA-125 measurements.

## 3. Results

### 3.1. Study cohort and assessment of technical variability

Plasma samples from 100 individuals, comprising 50 patients diagnosed with high-grade serous ovarian carcinoma (HGSOC) and 50 cancer-free controls, were initially included in the study. Samples were obtained from four biobanks: the Biobanco del Sistema de Salud de Aragón (BSSA), Biobanco de Navarra BioMed, Biobanco Vasco, and Biobanco para la Investigación Biomédica y en Salud Pública de la Comunidad Valenciana (IBSP-CV). Following quality-control procedures, 94 samples were retained for downstream analyses, resulting in a final analytical cohort of 47 patients with ovarian cancer and 47 controls. The mean age was comparable between groups (61.5 ± 12.3 years in patients and 62.2 ± 11.1 years in controls). According to the available FIGO classification, 2 patients had stage I disease, 3 had stage II, 36 had stage III, and 6 had stage IV. Accordingly, 5 patients had early-stage disease (FIGO I-II), whereas 42 presented with advanced-stage disease (FIGO III-IV). Additional information, including serum CA-125 concentrations, BRCA1/2 mutational status, and homologous recombination deficiency (HRD), was collected when available. Molecular testing was not uniformly performed across the cohort, and genetic findings were therefore considered descriptive. Demographic and clinicopathological characteristics are summarized in Table 1. The distribution of samples across biobanks was markedly unbalanced between study groups. Among controls, 45 of 47 samples originated from IBSP-CV and 2 from BSSA, whereas cancer samples originated from BSSA (n = 25), Navarra BioMed (n = 17), and the Biobanco Vasco (n = 5). Consequently, biobank of origin was strongly associated with disease status. Initial quality-control analyses revealed variability in the Ct values of the exogenous cel-miR-39-3p spike-in control across biobanks (Supplementary Fig. S1), indicating potential differences in sample processing or RNA recovery. This observation was considered when interpreting downstream expression analyses, as the substantial imbalance in biobank representation prevented reliable separation of disease-associated differences from potential pre-analytical effects.

**Table 1.** Demographic and clinicopathological characteristics of the study cohort. Continuous variables are presented as mean ± standard deviation (SD) or median and interquartile range (IQR), as appropriate, while categorical variables are expressed as absolute frequencies. FIGO stage was available for all 47 patients with HGSOC. Serum CA-125 concentrations were available for 17 patients from Navarra BioMed and are therefore reported exclusively for this subgroup. BRCA1/2 molecular results were available for seven patients, including two with confirmed pathogenic variants, one with a variant reported as likely pathogenic, and four with no pathogenic variants detected in the available testing. Homologous recombination deficiency (HRD) positivity was documented in one patient.

| **Characteristic** | **Controls (n = 47)** | **Cancer (n = 47)** |
|---|---|---|
| Age, years | 62.2 ± 11.1 | 61.5 ± 12.3 |
| High-grade serous carcinoma | - | 47 (100%) |
| **FIGO stage, n** |  |  |
| I | - | 2 |
| II | - | 3 |
| III | - | 36 |
| IV | - | 6 |
| **Molecular markers** |  |  |
| CA-125, kU/L, median (IQR) | - | 1,161 (254-3,635); n = 17 |
| BRCA1/2 pathogenic variant | - | 2 |
| BRCA1/2 likely pathogenic variant reported | - | 1 |
| BRCA1/2 no pathogenic variant detected | - | 4 |
| HRD positive confirmed | - | 1 |
| **Biobank, n** |  |  |
| BBSA | 2 | 25 |
| Navarra BioMed | 0 | 17 |
| BB Vasco | 0 | 5 |
| IBSP-CV | 45 | 0 |

### 3.2. Diagnostic performance of circulating hsa-miR-23a-3p

Circulating expression levels of eight candidate miRNAs (hsa-miR-23a-3p, hsa-miR-21-5p, hsa-miR-125b-5p, hsa-miR-93-5p, hsa-miR-20a-5p, hsa-miR-200c-3p, hsa-miR-200a-3p, and hsa-miR-320b) were evaluated in plasma samples from 47 patients with HGSOC and 47 controls. Among the investigated candidates, hsa-miR-23a-3p showed a significant difference in normalized circulating expression between patients and controls, with lower levels observed in the HGSOC group (Wilcoxon rank-sum test, p = 2.14 × 10^−5^; Fig. 1A). Expression profiles of the complete miRNA panel are presented in Supplementary Fig. S2. Receiver operating characteristic (ROC) curve analysis was performed to assess the discriminatory performance of circulating hsa-miR-23a-3p. The marker achieved an area under the curve (AUC) of 0.755 (95% CI, 0.657-0.852), indicating moderate discrimination between patients with HGSOC and controls (Fig. 1B). At the threshold selected using the Youden index, sensitivity was 93.6% and specificity was 53.2%. To explore the relationship between circulating hsa-miR-23a-3p expression and disease stage, normalized expression levels were examined across FIGO stages I-IV (Fig. 1C). The distribution of expression values was assessed in patients with stage I (n = 2), stage II (n = 3), stage III (n = 36), and stage IV (n = 6) disease. These analyses were considered exploratory because of the limited representation of early-stage tumors, particularly FIGO stage I. Accordingly, the available data do not allow definitive conclusions regarding the ability of circulating hsa-miR-23a-3p to discriminate early-stage from advanced-stage disease.

**Figure 1.**
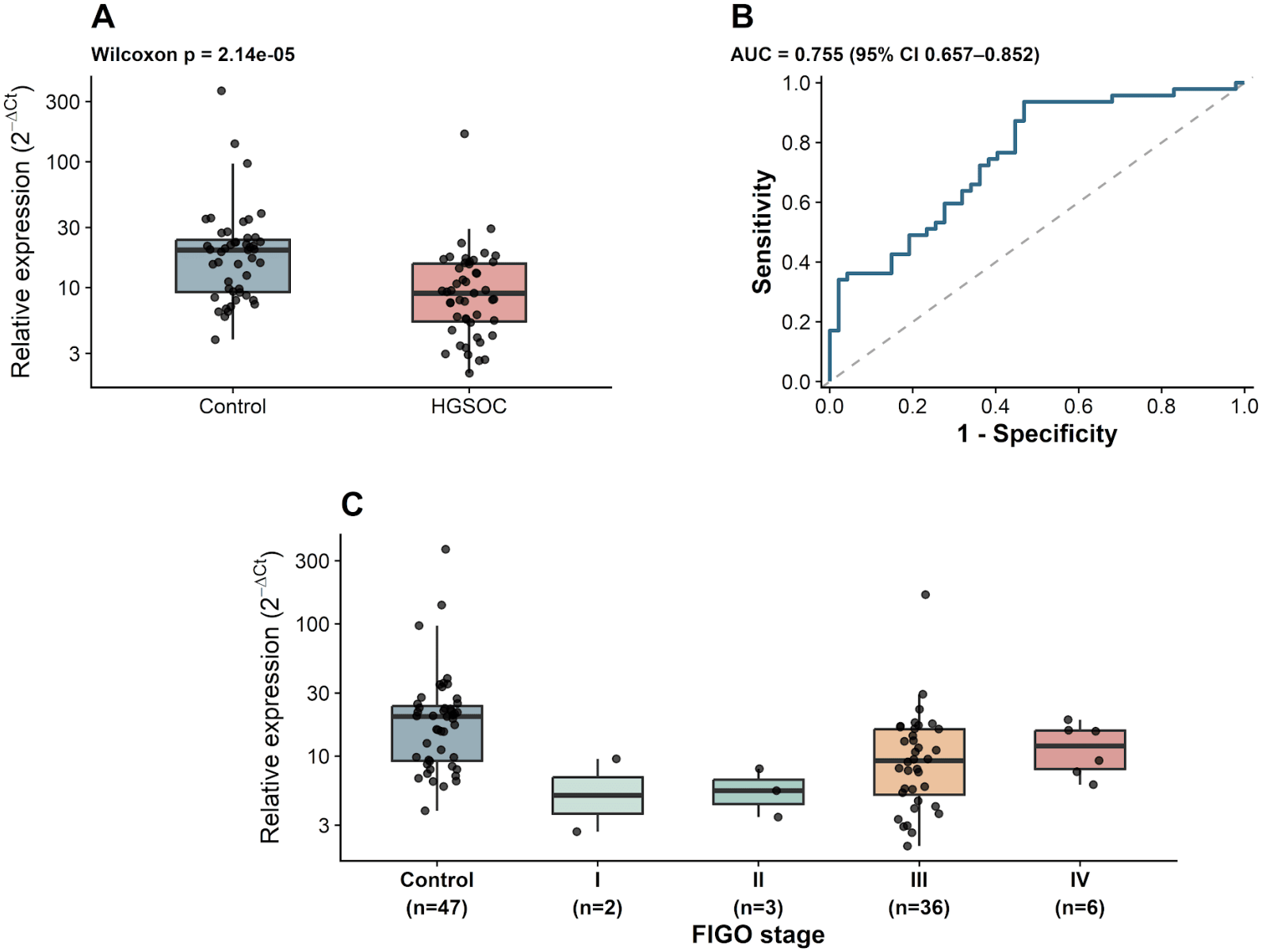
Circulating hsa-miR-23a-3p expression and discriminatory performance in patients with high-grade serous ovarian carcinoma (HGSOC). (A) Relative plasma expression of hsa-miR-23a-3p in healthy controls (n = 47) and patients with HGSOC (n = 47). (B) Receiver operating characteristic (ROC) curve evaluating the ability of circulating hsa-miR-23a-3p to discriminate patients with HGSOC from healthy controls. The area under the curve (AUC) was 0.755 (95% CI, 0.657-0.852). (C) Distribution of circulating hsa-miR-23a-3p expression according to FIGO stage: stage I (n = 2), stage II (n = 3), stage III (n = 36), and stage IV (n = 6), compared with healthy controls (n = 47). Relative expression was calculated as 2^-ΔCt^ following global mean normalization and is displayed on a logarithmic scale. Boxplots represent the median and interquartile range, with whiskers extending to 1.5× the interquartile range; individual dots represent individual participants. The comparison between controls and the overall HGSOC group was performed using the Wilcoxon rank-sum test. CI, confidence interval; FIGO, International Federation of Gynecology and Obstetrics; HGSOC, high-grade serous carcinoma; ROC, receiver operating characteristic.

### 3.3. Exploratory evaluation and internal validation of a three-miRNA circulating signature

To investigate whether combining circulating miRNAs could improve discrimination between patients with HGSOC and controls, all 56 possible three-miRNA combinations were evaluated using multivariable logistic regression models. The combination of hsa-miR-23a-3p, hsa-miR-21-5p, and hsa-miR-20a-5p achieved the highest apparent discriminatory performance, with an area under the receiver operating characteristic curve (AUC) of 0.798 (Fig. 2A), compared with 0.755 for hsa-miR-23a-3p alone (Fig. 2B). To evaluate the generalizability of this approach and account for potential model-selection optimism, repeated stratified five-fold cross-validation was performed over 20 repetitions, with the selection of the three-miRNA combination repeated within each training partition. The mean cross-validated AUC was 0.716 ± 0.022, indicating a reduction in discriminatory performance compared with the apparent AUC estimated from the original dataset. Although the three-miRNA signature showed numerically higher apparent discrimination than hsa-miR-23a-3p alone, its performance was attenuated following internal validation, suggesting that the initial improvement may partly reflect model-selection optimism. Furthermore, the marked imbalance in biobank representation between study groups introduces potential pre-analytical confounding. Therefore, these findings should be considered exploratory, and independent validation in appropriately balanced cohorts is required to establish the clinical relevance of the proposed signature.

**Figure 2.**
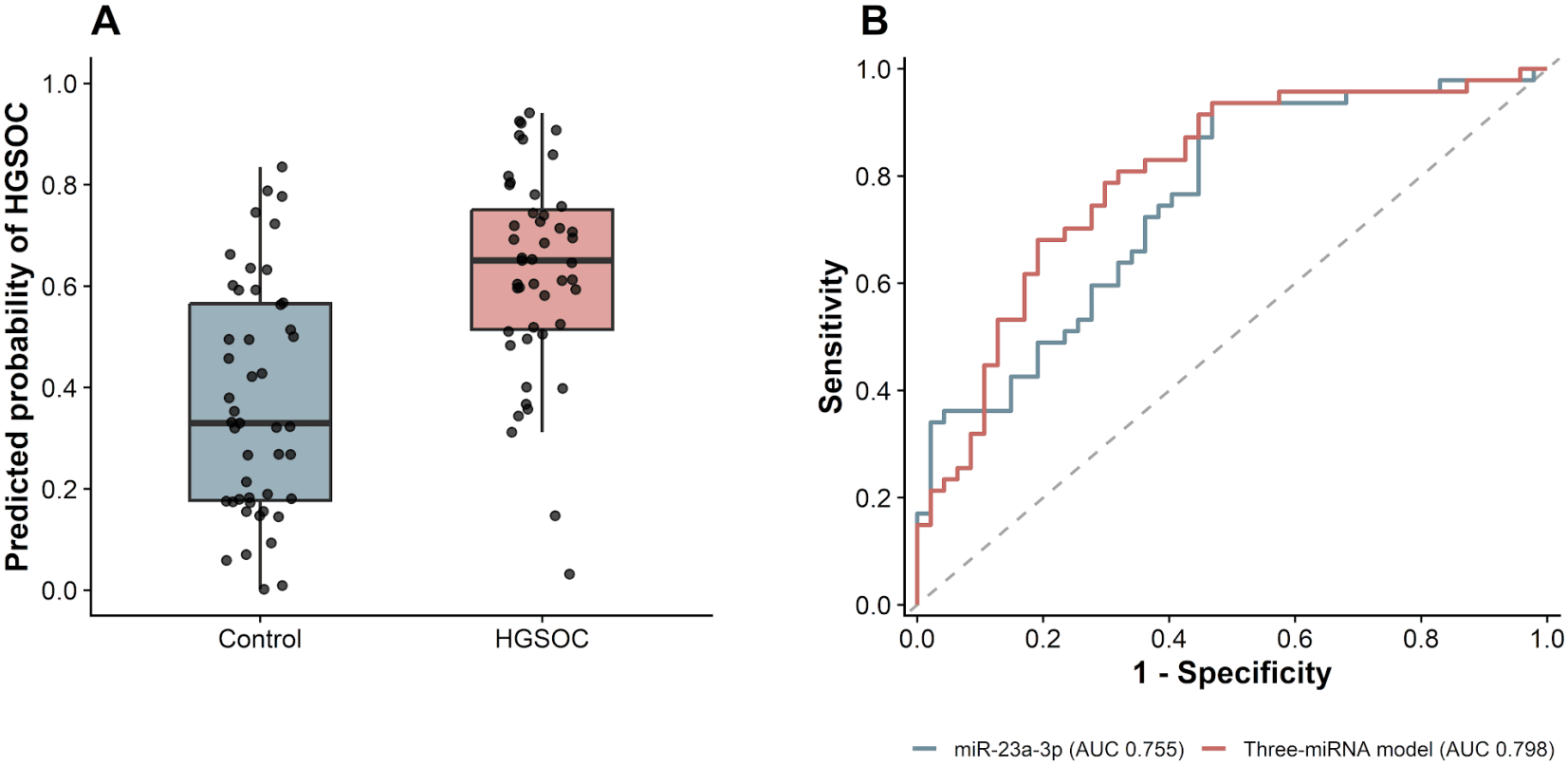
Exploratory discriminatory performance of a three-miRNA circulating signature in high-grade serous ovarian carcinoma (HGSOC). (A) Distribution of predicted HGSOC probabilities obtained from a logistic regression model incorporating hsa-miR-23a-3p, hsa-miR-21-5p, and hsa-miR-20a-5p in controls (n = 47) and patients with HGSOC (n = 47). Boxplots represent the median and interquartile range, with whiskers extending to 1.5 times the interquartile range; individual dots represent plasma samples. (B) Receiver operating characteristic (ROC) curves comparing hsa-miR-23a-3p alone (apparent AUC = 0.755) and the three-miRNA signature (apparent AUC = 0.798). Both ROC curves represent apparent discriminatory performance estimated using the model-development dataset. Repeated stratified five-fold cross-validation (20 repetitions), with miRNA selection performed within each training partition, yielded a mean AUC of 0.716 ± 0.022 for thethree-miRNA model-selection strategy. Cross-validated performance is not depicted in the ROC curves

### 3.4. Association between circulating hsa-miR-23a-3p expression and serum CA-125 levels

In the subset of 17 patients with HGSOC for whom serum CA-125 measurements were available, an exploratory Spearman correlation analysis was performed to assess the relationship between circulating hsa-miR-23a-3p expression and CA-125 concentrations. A positive but non-statistically significant correlation was observed (Spearman.s ρ = 0.301, p = 0.240; Supplementary Fig. S3). These findings did not provide evidence of a significant association between circulating hsa-miR-23a-3p expression and serum CA-125 levels in this limited subgroup.

## Discussion

In the present study, we investigated the circulating expression profiles of eight candidate miRNAs in plasma samples from patients with high-grade serous ovarian carcinoma (HGSOC) and cancer-free controls. Among the evaluated candidates, hsa-miR-23a-3p exhibited significantly lower relative expression in patients with HGSOC and achieved moderate discriminatory performance, with an AUC of 0.755. Furthermore, an exploratory three-miRNA signature incorporating hsa-miR-23a-3p, hsa-miR-21-5p, and hsa-miR-20a-5p achieved an apparent AUC of 0.798, although its performance decreased following internal cross-validation. These findings support further investigation of circulating miRNA profiles as potential biomarkers in HGSOC while emphasizing the methodological challenges associated with their clinical translation.

The findings reported are consistent with previous investigations reporting disease-associated alterations in circulating miRNAs, despite differences in the specific biomarkers evaluated. Elevated circulating levels of miR-200a, miR-200b, and miR-200c have been reported in patients with HGSOC compared with healthy women, with the combination of miR-200b and miR-200c proposed as a potential diagnostic signature [21]. Similarly, the combined evaluation of miR-205 and miRNA let-7f has been investigated for distinguishing epithelial ovarian cancer patients from healthy controls, particularly in early-stage disease [22]. Other studies have identified miR-1246, miR-595, and miR-2278 as potential circulating biomarkers for differentiating HGSOC from healthy individuals, with findings supported by independent discovery and validation cohorts [23]. In addition, exosomal miR-1290 has been investigated for discriminating HGSOC patients from healthy controls and patients with ovarian malignancies of other histological subtypes [24]. Similarly, the combination of miR-200c-3p and miR-221-3p has been proposed for differentiating malignant from benign pelvic masses, with its diagnostic potential further supported by validation in an independent cohort [25]. More recently, a distinct serum signature comprising miR-1246 and miR-150-5p was identified for distinguishing HGSOC patients from healthy women [26]. Collectively, these findings support the diagnostic potential of circulating miRNAs, while highlighting the substantial heterogeneity in the molecular signatures reported across investigations.

Beyond individual miRNAs and small biomarker combinations, other studies have explored more complex miRNA panels or focused on different clinical comparisons. A seven-miRNA plasma signature comprising miR-92a, miR-200c, miR-320b, miR-320c, miR-335, miR-375, and miR-486 has been evaluated for distinguishing ovarian cancer patients and women with benign ovarian lesions from healthy controls [27]. Larger serum-based profiling studies have also identified a ten-miRNA signature for discriminating ovarian cancer patients from non-cancer controls, while additional panels comprising nine or ten miRNAs were developed to assess discrimination from other malignancies and benign ovarian tumors [28]. Furthermore, the eight-miRNA OCaMIR signature, comprising miR-182, miR-183, miR-202, miR-205, miR-508, miR-509-3, miR-513b, and miR-513c, was developed through molecular profiling of early-stage serous ovarian cancer tissue and subsequently evaluated in independent retrospective and prospective serum cohorts [29]. These investigations illustrate that the reported molecular signatures differ not only in their miRNA composition but also in their intended clinical applications, ranging from the discrimination of HGSOC patients from healthy individuals to the differentiation of malignant and benign ovarian conditions or the identification of early-stage disease. Consequently, discrepancies between studies may reflect differences in comparator populations, histological composition, biological matrices, and analytical methodologies, rather than necessarily indicating inconsistent biological findings.

Despite the promising diagnostic performance reported for multivariable miRNA signatures, their translation into routine clinical practice remains challenging. The simultaneous quantification of multiple targets may increase analytical complexity and place additional demands on assay standardization, quality control, and model validation [30,31]. In this context, the identification of a single circulating miRNA showing discriminatory potential, as observed for hsa-miR-23a-3p in the present study, may offer practical advantages by facilitating the development of a simpler and potentially more cost-effective targeted assay. Nevertheless, analytical simplicity does not necessarily imply superior clinical utility, particularly given the generally improved diagnostic performance reported for multivariable signatures in meta-analytical investigations [13]. From a biological perspective, miR-23a-3p has been implicated in several processes relevant to ovarian cancer progression, including the regulation of cell proliferation, apoptosis, migration, and invasion [32,33]. However, its functional effects are context-dependent, and circulating miRNA abundance may reflect multiple cellular sources and mechanisms of extracellular release rather than directly representing intratumoral expression or biological activity [30,32]. Interestingly, hsa-miR-23a-3p has also been proposed as a stable endogenous reference in plasma from HGSOC patients and women with benign ovarian tumors [34]. However, the absence of healthy controls in that investigation limits the applicability of its findings to the present comparison. Thus, the lower relative circulating expression observed in our cohort does not necessarily contradict its previously reported stability in different clinical populations, but reinforces the importance of evaluating miRNA behavior within the specific biological and analytical context under investigation.

A relevant feature of the present study is the evaluation of circulating hsa-miR-23a-3p expression according to FIGO stage within a histologically homogeneous HGSOC cohort. Although previous studies have considered disease stage, many investigations have combined different ovarian cancer histological subtypes or have not performed detailed stage-stratified analyses. In this context, our approach provides a more specific assessment of miRNA expression in relation to disease extent. Nevertheless, the limited representation of early-stage patients (n = 5, FIGO I-II) precludes definitive conclusions regarding its potential for early detection. Additionally, no significant correlation was observed between hsa-miR-23a-3p expression and CA-125 concentrations (Spearman.s ρ=0.301, p=0.240; n=17), although the limited sample size prevents firm conclusions regarding their relationship or potential diagnostic complementarity. A further methodological consideration is the inclusion of plasma obtained from multiple independent biobanks. This approach introduces additional preanalytical heterogeneity, potentially reflecting differences in blood collection procedures, processing intervals, centrifugation protocols, storage duration, and sample handling. These factors have been demonstrated to influence circulating miRNA recovery and quantification and may contribute to variability between biomarker studies [35,36]. Although synthetic spike-in controls are useful for monitoring technical variation during RNA extraction and downstream amplification, they cannot account for differences introduced before their addition, including blood processing delays, cellular contamination, or storage-related changes [36,37]. Despite this heterogeneity, hsa-miR-23a-3p exhibited significantly different circulating expression between HGSOC patients and controls, supporting its potential as a candidate biomarker. Although further validation is required to disentangle disease-associated differences from biobank-related effects, the inclusion of samples from diverse sources provides a valuable basis for investigating the reproducibility and clinical applicability of this miRNA under heterogeneous preanalytical conditions.

The development of reliable, minimally invasive biomarkers remains a major priority in ovarian cancer research, particularly given the challenges associated with early diagnosis and the limited specificity of currently available diagnostic approaches. In this context, circulating miRNAs represent promising candidates for improving disease detection and patient stratification, potentially complementing conventional clinical and molecular assessments. However, their successful translation into clinical practice requires further validation of their diagnostic performance, reproducibility, and applicability across heterogeneous patient populations. Future investigations should therefore focus on validating hsa-miR-23a-3p and predefined multivariable signatures in larger, prospectively collected cohorts, incorporating different FIGO stages, clinically relevant control groups, and standardized preanalytical procedures. Furthermore, integrating circulating miRNA measurements with established biomarkers, particularly CA-125, molecular characteristics, and longitudinal clinical information may help clarify their potential added diagnostic value and relevance for disease monitoring. Overall, the present findings identify hsa-miR-23a-3p as a promising candidate associated with altered circulating expression in HGSOC, providing a basis for further investigations aimed at establishing its potential clinical utility.

## 5. Conclusion

This exploratory study identified significantly lower normalized circulating hsa-miR-23a-3p expression in patients with high-grade serous ovarian carcinoma compared with cancer-free controls, with moderate apparent discriminatory performance (AUC = 0.755). A three-miRNA signature incorporating hsa-miR-23a-3p, hsa-miR-21-5p, and hsa-miR-20a-5p achieved a higher apparent AUC of 0.798; however, its performance decreased following repeated cross-validation (mean AUC = 0.716 ± 0.022), indicating potential model-selection optimism. No statistically significant association was observed between circulating hsa-miR-23a-3p expression and serum CA-125 concentrations in the limited subgroup with available clinical data. Importantly, the pronounced imbalance in biobank representation between patients and controls represents a major source of potential pre-analytical confounding and limits the attribution of the observed expression differences to disease status. Therefore, these findings should be considered hypothesis-generating. Independent validation in larger, prospectively collected cohorts with harmonized sample processing and balanced recruitment is essential to determine the biological relevance and potential clinical utility of circulating hsa-miR-23a-3p and multivariable miRNA signatures in HGSOC.

## Data Availability

All data produced in the present study are available upon reasonable request to the authors

## 7. Author Contributions

MEO, GGL and ELG performed the analyses and interpreted the results; AMT and ASR critically reviewed and revised the manuscript; AL conceived the original idea and critically reviewed the manuscript; MEO drafted the original version of the manuscript and served as the corresponding author. All authors read and approved the final version of the manuscript.

## 9. Acknowledgments

The authors gratefully acknowledge the participating biobanks and their staff for their valuable collaboration in providing the biological samples used in this study.

## 10. Institutional Review Board Statement

The study was approved by the Public Health Research Ethics Committee (CEI-SP) of the Foundation for the Promotion of Health and Biomedical Research of the Valencian Community (FISABIO), Spain (approval no. 20250717/06/P; 17 July 2025), in accordance with the Declaration of Helsinki and applicable Spanish and European regulations.

## 11. Informed Consent Statement

Participants or their family members/legal guardians provided written informed consent for the collection, storage, and research use of biological samples, with consent documentation managed and retained by the participating biobanks.

