## Supplemental File for "Circulating miRNA profiling identifies hsa-miR-23a-3p and a three-miRNA signature associated with serous ovarian cancer"

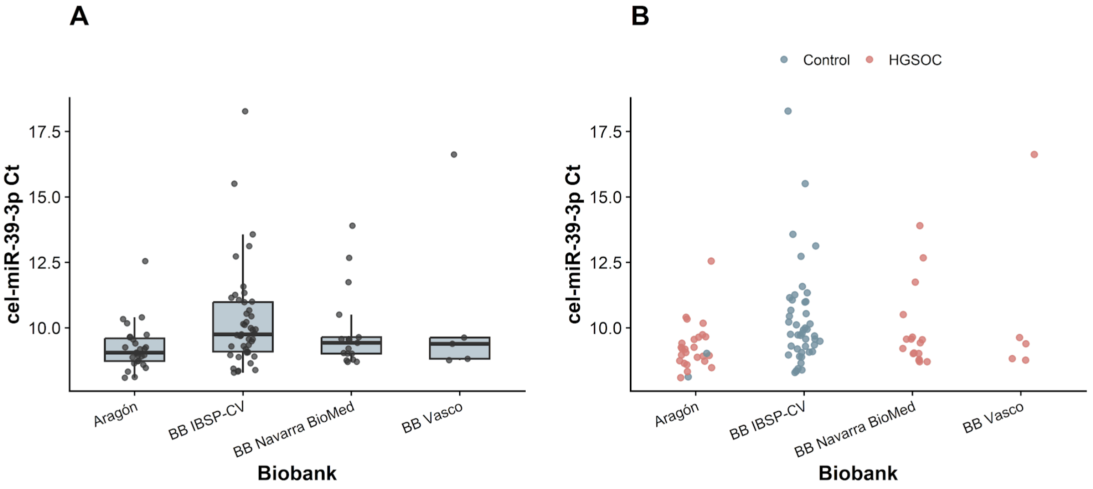


**Supplementary Figure S1. Inter-biobank variability in the exogenous cel-miR-39-3p spike-in control.** (A) Distribution of cel-miR-39-3p cycle threshold (Ct) values across the four participating biobanks: Aragón, IBSP-CV, Navarra BioMed, and the Basque Biobank. Boxplots represent the median and interquartile range (IQR), with whiskers extending to 1.5 times the IQR. Individual dots represent plasma samples. (B) Distribution of cel-miR-39-3p Ct values according to biobank of origin and study group (controls and HGSOC). Greater variability was observed in samples from IBSP-CV, whereas the remaining biobanks showed comparatively narrower distributions. The marked imbalance in sample provenance between study groups highlights the potential influence of pre-analytical and technical variability on circulating miRNA measurements.

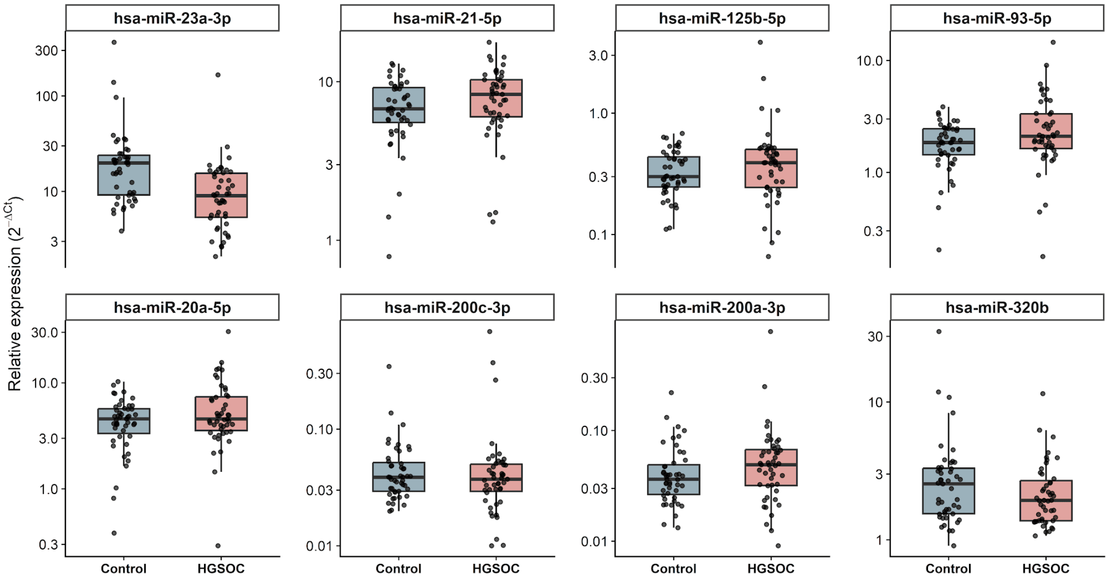


**Supplementary Figure S2. Circulating expression profiles of eight candidate miRNAs in patients with high-grade serous ovarian carcinoma (HGSOC) and controls.** Relative plasma expression levels of hsa-miR-23a-3p, hsa-miR-21-5p, hsa-miR-125b-5p, hsa-miR-93-5p, hsa-miR-20a-5p, hsa-miR-200c-3p, hsa-miR-200a-3p, and hsa-miR-320b in controls (n = 47) and patients with HGSOC (n = 47). Relative expression was calculated as $2^{-\Delta Ct}$ following global mean normalization and is displayed on a logarithmic scale. Boxplots represent the median and interquartile range (IQR), with whiskers extending to 1.5 times the IQR. Individual dots represent individual plasma samples.

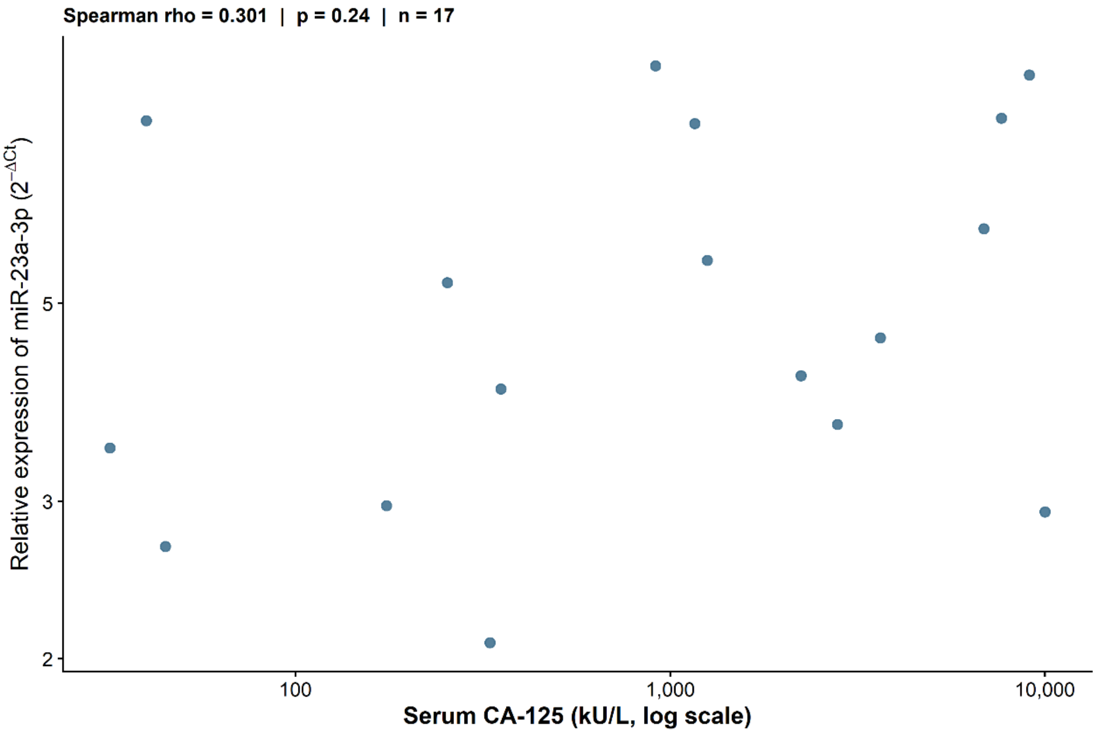


**Supplementary Figure S3. Exploratory correlation between circulating hsa-miR-23a-3p expression and serum CA-125 concentrations in patients with HGSOC.** Scatter plot showing relative hsa-miR-23a-3p expression ($2^{-\Delta Ct}$) against serum CA-125 concentrations (kU/L, logarithmic scale) in 17 patients. Each point represents an individual patient. Spearman's rank correlation analysis showed no statistically significant association ($\rho = 0.301$, $p = 0.240$).
