## Supplementary figures and images for "Circulating miRNA profiling identifies hsa-miR-23a-3p and a three-miRNA signature associated with serous ovarian cancer"

### Supplementary_Figure_S1

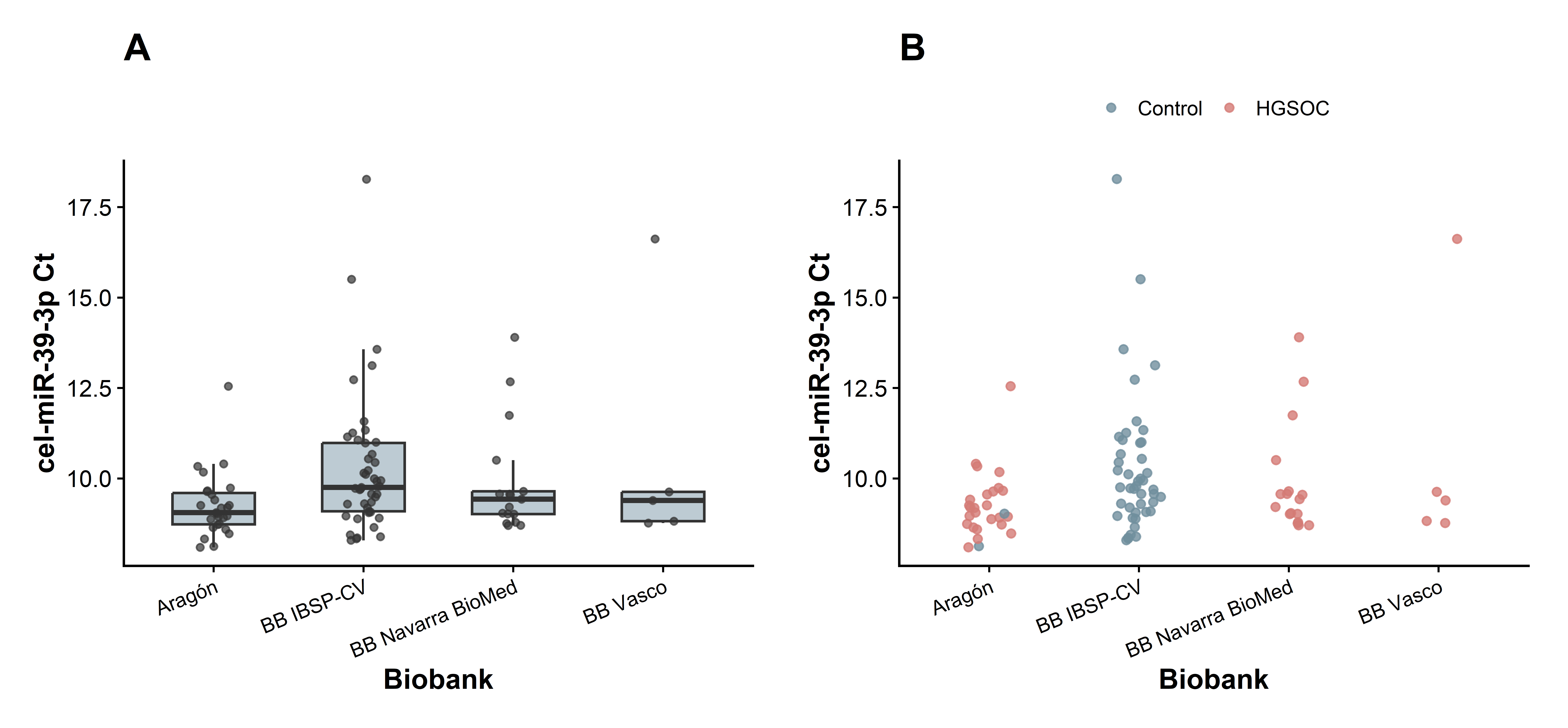

### Supplementary_Figure_S2

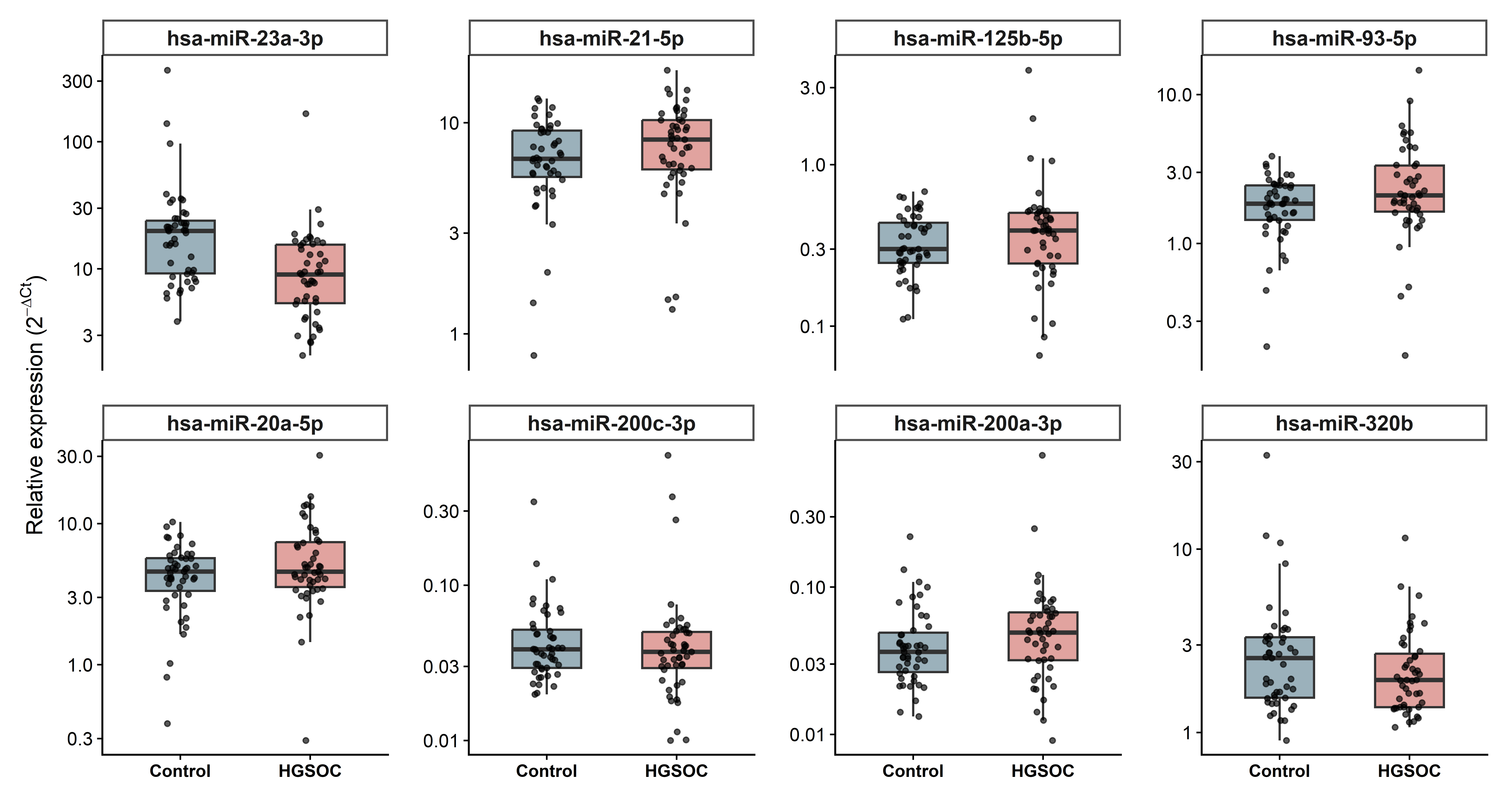

### Supplementary_Figure_S3

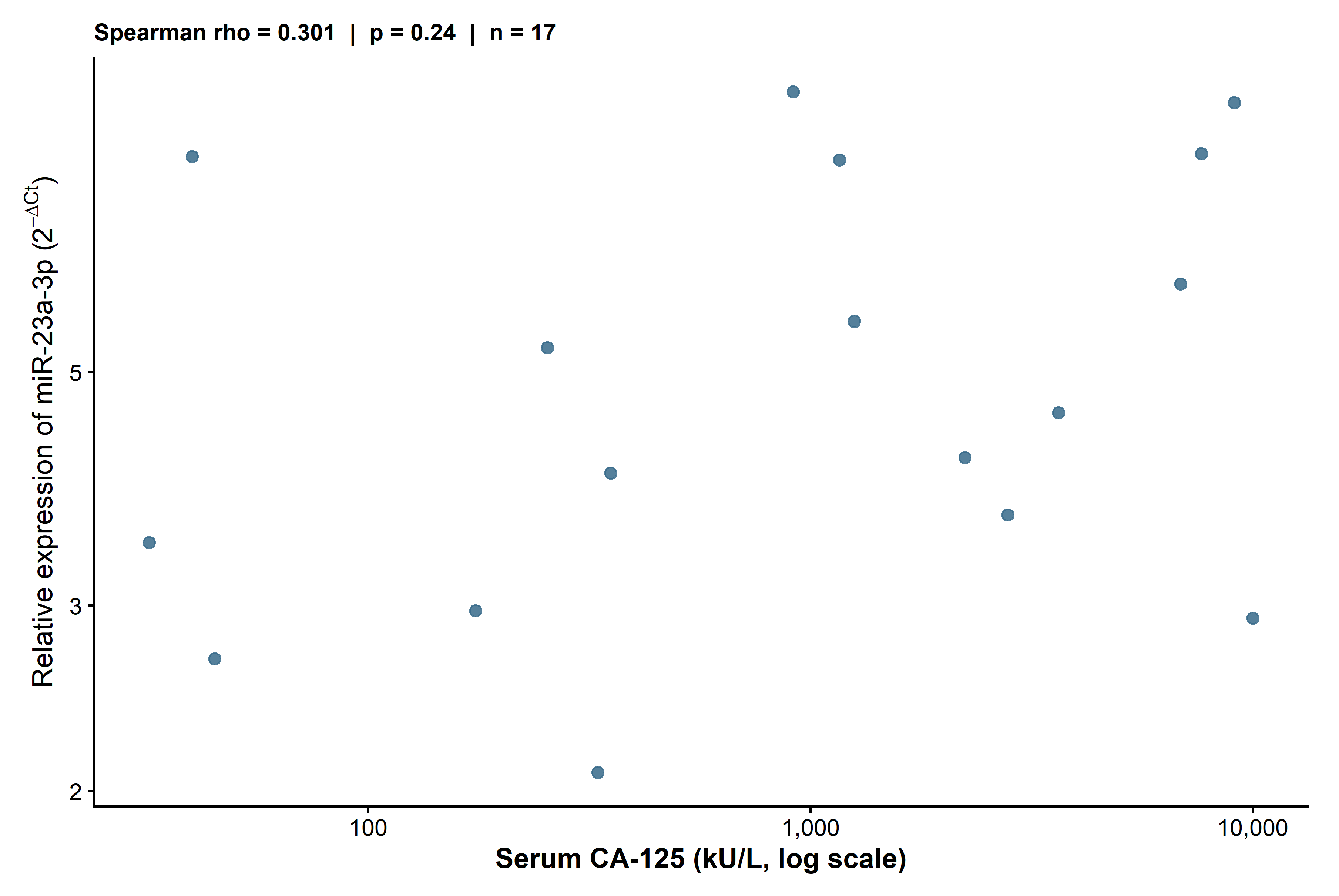
